# Combing the haystacks: The search for highly pathogenic avian influenza virus using a combined clinical and research-developed testing strategy

**DOI:** 10.1101/2025.02.12.25321810

**Authors:** Gordon C. Adams, Jamie E. Devlin, Erik Klontz, Rachel A. Laing, John A. Branda, Navid Chowdhury, SunYoung Kwon, Pardis C. Sabeti, Elyse Stachler, Vamsi Thiriveedhi, Erica S. Shenoy, Jacob E Lemieux, Sarah E Turbett

## Abstract

**Background:** Highly pathogenic H5 avian influenza A has caused sporadic human infections, increasing the risk for potential human-to-human spread. In 2024, the U.S. experienced outbreaks among poultry and cattle, prompting enhanced surveillance.

**Objective:** To evaluate an H5 testing algorithm in subjects with respiratory symptoms presenting for routine care during low influenza A virus circulation.

**Design:** Observational study using clinical- and research-developed nucleic acid amplification tests (NAATs) and pooled screening methods.

**Setting:** Academic medical center in Boston, MA.

**Participants:** 5,400 symptomatic individuals contributing 6,935 respiratory specimens from June 23 to August 28, 2024.

**Measurements:** Specimens underwent initial respiratory pathogen testing per clinical protocols, which did not routinely include influenza due to low summer-month prevalence. Influenza A-positive specimens were subtyped using a clinical assay for H5 assessment. SARS-CoV-2-negative specimens not tested for influenza were screened in pooled batches. Positive pools were deconvoluted to individual specimens and screened for H5 using quantitative polymerase chain reaction.

**Results:** Influenza A was detected in 40 of 6,935 specimens (0.6%), comprising 35 of 5,400 unique subjects (0.7%). No H5 infections were identified. Of the 35 influenza-positive individuals, 10 cases (29%) were found through research-specific screening of SARS-CoV-2–negative specimens. No deaths attributed to influenza were recorded.

**Limitations:** Single center design, convenience sampling, absence of ocular specimens, and minimal sampling in high-risk areas may limit generalizability.

**Conclusion:** Expanded influenza testing using pooled NAATs successfully identified low-prevalence influenza A and ruled out H5 in this cohort. These data support targeted influenza screening to enhance surveillance for emerging subtypes rather than a broad-based clinical testing strategy for influenza A testing.

**Primary Funding Source:** Supported by institutional resources and Center for Disease Control and Prevention contracts.

## Background

Since the identification of highly pathogenic avian influenza A virus H5 (H5) in 1996, the virus has primarily circulated in birds, with endemic rates throughout southeastern Asia since 2009 (1). Human H5 infections have occurred sporadically, with the first cases confirmed in 18 individuals in Hong Kong in 1997 (2). These infections, acquired from chickens, caused death in one third of cases (2). Since then, nearly 1,000 human infections have been reported worldwide; human-to-human transmission has remained limited, with the last reported case occurring in 2007 (3). In the U.S., human cases were first identified in 2022 amid a poultry outbreak linked to animal exposure (4,5). This outbreak expanded to dairy cattle, with herd infections spanning multiple states (6,7). As of January 31, 2025, 67 human cases have been linked to the overall outbreak, with the first death from severe disease in January of 2025 (8). To date, there has been no evidence of sustained or non-sustained human-to-human transmission (7).

In response to the widespread poultry and cattle outbreak, the United States Centers for Disease Control and Prevention (CDC) recommended clinicians continue testing all individuals with respiratory symptoms for influenza A during the summer of 2024, instead of usual guidance to only test for influenza A when the virus is circulating at moderate-to-high levels, and only when the test result will change clinical management (9–13). This recommendation was made to strengthen H5 surveillance and to detect cryptic cases in clinical settings, particularly when influenza A testing is often limited by clinical laboratories based on existing guidance and low disease prevalence.

In response to the updated CDC guidance, a custom-designed algorithm was developed integrating both clinical laboratory and research-developed nucleic acid amplification tests (NAATs) to detect H5 in symptomatic subjects undergoing respiratory virus testing at hospital-based clinical laboratories at Massachusetts General Hospital (MGH). Here, we describe the implementation of our novel algorithm and describe its findings.

## Methods

### Sample collection and H5 testing algorithm

Respiratory specimens were tested for H5 from symptomatic persons presenting for clinical care at MGH and its affiliated outpatient practices using a custom-designed algorithm integrating clinical laboratory and research-developed NAATs (Figure 1). Specimens were initially tested for respiratory pathogens using one of the following assays: Xpert Xpress CoV-2 Plus (CoV-2 Plus, Cepheid, Sunnyvale, CA), Xpert Xpress CoV-2/Flu/RSV Plus (CoV-2/Flu/RSV Plus, Cepheid, Sunnyvale, CA), or the BioFire Respiratory 2.1 Panel (RP2.1, bioMerieux, Salt Lake City, UT). The CoV-2 Plus tests for SARS-CoV-2; the CoV-2/FLU/RSV Plus tests for SARS-CoV-2, influenza A and B, and respiratory syncytial virus (RSV); and the RP 2.1 tests for the following viruses: SARS-CoV-2, non-SARS-CoV-2 coronaviruses, human rhinovirus/enterovirus, influenza A and B, RSV, parainfluenza viruses 1-4, adenovirus, and human metapneumovirus. All assays are Food and Drug Administration (FDA) emergency use authorized or cleared and clinically verified in the MGH Clinical Microbiology Laboratory. In the outpatient setting, test choice was determined through an order set that incorporates best practice guidance for respiratory virus testing (14). In the emergency department and inpatient settings, test choice was determined using embedded clinical decision support that combines clinical and operational questions to determine which targets will be evaluated, which was vetted by clinical leadership, subject matter experts, and supported by national guidelines.

**Figure 1.**
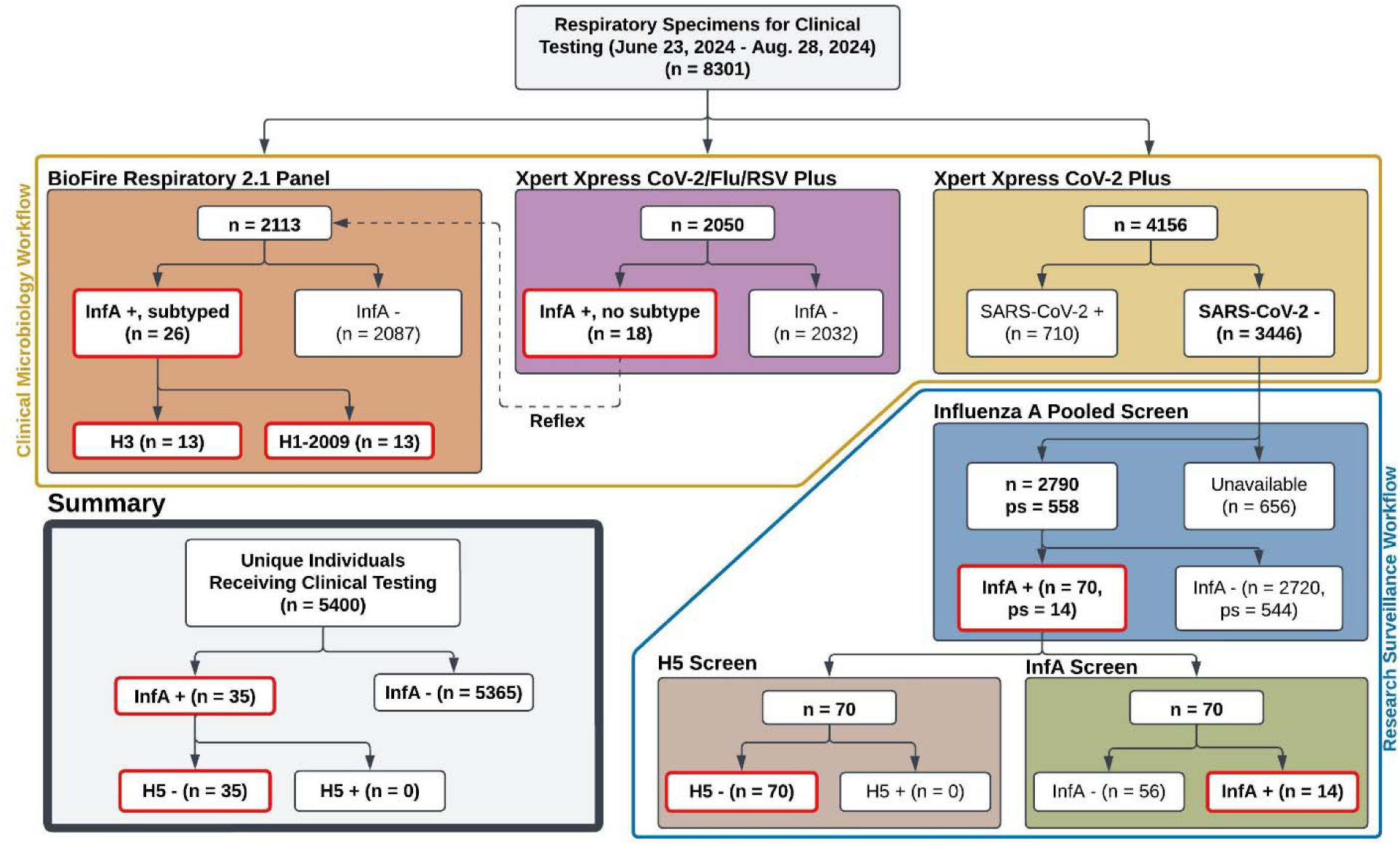
Clinical and research NAAT Pan-Influenza A and H5 testing workflow for respiratory specimens collected between 06/23/24 and 8/28/24 from patients seeking care at MGH and its affiliated outpatient practices. Unique specimen numbers are reported using “n” and pools of 5 unique specimens are reported as “ps”. Testing conducted through the adapted clinical microbiology workflow is outlined in yellow, and screening through research surveillance methods is outlined in blue. Key findings are highlighted in red. A total of 40 specimens from 35 unique individuals were positive for influenza A during the collection period: 26 identified and typed through clinical testing, and 14 identified through the research surveillance workflow. No specimens were found to be positive for H5. InfA: influenza A; “+”: positive; “-”: negative; NAAT: nucleic acid amplification testing; MGH: Massachusetts General Hospital

The CoV-2/FLU/RSV Plus detects segments of the matrix, polymerase, and acidic protein of the influenza A genome; it can detect H5 but cannot differentiate it from other influenza A subtypes In contrast, the RP 2.1 detects genomic segments present in all influenza A subtypes, including H5, as well as common seasonal influenza hemagglutinin subtypes (H1, H3, H1-2009) Novel influenza A subtypes, including H5, are reported as “influenza A-no subtype detected”. During the collection period, any specimen reported as “influenza A-no subtype detected” was retested on the RP 2.1 per the manufacturer’s recommendations. Were a repeat result of “Influenza A-no subtype detected” identified, the Massachusetts Department of Public Health (MADPH) would be notified for consideration of H5 confirmatory testing.

During the collection period, select individuals underwent only SARS-CoV-2 testing using the algorithm described above. Those who tested negative for SARS-CoV-2 were eligible for influenza A screening using a pooled specimen testing strategy with an internally developed pan-influenza A NAAT. Any pool that tested positive was subsequently deconvoluted, and each specimen within that pool was tested with both pan-influenza A and H5-specific NAATs to assess for individual H5 cases (Figure 1).

### Influenza A and H5 nucleic acid amplification assay design

Influenza A screening utilized the CDC’s Influenza and SARS-CoV-2 Multiplex assay with the following modifications: the SARS-CoV-2 and influenza B primers and probes were removed; and two degeneracy points were introduced into one of the influenza A reverse primers based on sequence analysis of publicly available H5N1 influenza A genomes isolated from cattle, flocks, or milk in the United States and published in National Center for Biotechnology (NCBI) databases (Supplement Table 1, (17). Other influenza A primers and the probe were unaltered, and the RNase P primers and probes were retained as an internal positive control (IPC). H5 screening was performed using strain specific primers and probes published previously (18).

Both the modified influenza A and H5 NAATs were validated using clinical specimens, artificial controls (DNA plasmids; Twist Biosciences, San Francisco, CA), and RNA from H5N1 positive cattle milk (19). I*n silico* analysis through Geneious prime was also performed to ensure detection of H5N1 clade 2.3.4.4b (Supplement Table 6). Sensitivity and specificity were assessed based on standard curves generated from synthetic control serial dilutions, and by testing both assays against BioFire-typed influenza A-H1N1-2009 and –H3N2, influenza B-positive clinical specimens, and influenza A H5N1 RNA isolated from milk (Supplementary Figure 1, Supplementary Table 3).

### Pooled screening of SARS-CoV-2 negative specimens

Influenza A and H5 screening of SARS-CoV-2 negative specimens was performed in two phases. First, five-specimen pools were created from randomly selected eligible respiratory specimens per FDA SARS-CoV-2 pooled specimen guidance, and tested for influenza A using the Pan-influenza NAAT (Supplemental Methods)(20,21). For the second phase, if a specimen pool tested positive or inconclusive for influenza A, individual specimens from that pool were re-extracted and screened for influenza A and H5 separately (in parallel) using the influenza A and H5 NAATs.

All qPCR analyses were performed using the ThermoFisher Cloud Analysis platform (ThermoFisher Scientific, Waltham, MA). Pools or specimens without a positive IPC signal were retested. All screening batches included positive and negative controls.

### Processing and analysis of geographical statistical data of respiratory specimens

To provide a geospatial representation of influenza A and H5 testing in our specimen cohort, patient identifiers and demographics, including medical record number (MRN), primary residence ZIP code, and specimen identification numbers, were obtained from all unique specimens through queries to clinical laboratory structured query language (SQL) databases. For individuals with primary residence in New England, testing counts were aggregated to the ZIP code level. ZIP code geometries were obtained from 2019 ZCTA5 shapefiles from U.S. Census Bureau data and ZIP-level population estimates were found via Simple Maps (22,23). Geographical metadata for a subset of ZIP codes had to be manually populated (Supplementary Table 7). Geospatial data were then merged with testing counts via the ZIP code and converted to rates per 100,000 persons. To correlate our testing catchment area with known flock or herd H5 outbreaks in New England, counties with confirmed flock or herd cases were obtained from the U.S. Department of Agriculture (USDA) and integrated into the geospatial data using county borders obtained from the U.S. Census Bureau (24).

### Clinical review of influenza A-positive individuals

Medical records of influenza A-positive subjects, identified either through clinical workflows or the research-specific screening process, were reviewed to gather demographics, clinical manifestations, comorbidities, disease course, and other relevant information. Each record was initially independently reviewed by two of four physicians (ESS, SET, EHK, JEL); any discordance was resolved by a third physician’s review. All data were obtained from the Mass General Brigham (MGB) electronic medical records with approval from the MGB Institutional Review Board (Protocol 2019P003305).

## Results

### Influenza A and H5 testing results

Between June 23 and August 28, 2024, 8,301 respiratory specimens were tested for respiratory pathogens in the MGH Clinical Microbiology Laboratory (Figure 1). Of these, 4,145 specimens (49.9%) underwent standard clinical laboratory influenza A NAAT, resulting in 26 positive tests (0.6%) from 25 unique individuals. All clinical positives were analyzed using an assay that provides subtypes (RP 2.1) which identified 13 (50%) as H3 and 13 (50%) as H1-2009 in 13 and 12 unique individuals respectively. No ambiguous H-type results were detected among clinically tested specimens.

The remaining 4,156 specimens (50.1%) were tested solely for SARS-CoV-2 (Figure 1), of which 710 (17.1%) were positive. The remaining 3,446 SARS-CoV-2–negative specimens (82.9%) were eligible for Influenza A and H5-specific screening using our research-developed workflow. Of these, 656 (19.0%) collected between July 24 and July 30, 2024 were unavailable for further testing. The remaining 2,790 specimens (67.1%) were screened using a pooled screening approach. First, specimens were tested using our influenza A NAAT in 558 5-specimen pools. Of those pools, 14 (2.5%) tested positive for influenza A. Individual specimens from each positive pool were subsequently screened, which identified 14 positive specimens (0.5%) from 10 unique individuals. None were positive for H5. In total, 6,935 specimens from 5,400 individuals were tested for influenza A through either clinical or research workflows, yielding 40 positive specimens (0.6%) from 35 individuals (0.7%).

### Geographic distribution of respiratory specimen collection

ZIP-level testing rates per 100,000 persons from symptomatic individuals in New England are described (Figure 2). In total, 4,875 (90.3%) individuals tested had primary residence in Massachusetts, 187 (3.5%) in New Hampshire, 60 (1.1%) in Maine, 48 (0.9%) in Rhode Island, 17 (0.3%) in Connecticut, and 16 (0.3%) in Vermont. There were 165 (3.0%) individuals with primary residence in the remaining 44 states, 29 (0.5%) individuals from outside of the U.S., and 3 individuals (<0.1%) for whom location data were unknown. Barnstable and Essex counties in MA and Knox, Kennebec, and York counties in ME, where H5 was detected in flocks within the preceding 6 months of the testing period, are outlined in red (Figure 2). Further outbreak details as reported by USDA APHIS are available in Supplementary Table 8. There were no H5 positive herds in New England during the testing period or in the preceding 6 months. In total, 577 individuals (10.7%) tested had primary residence in Essex County, 44 (0.8%) in Barnstable County, 1 (<0.1%) in Knox County, 6 (0.1%) in Kennebec County, and 11 (0.2%) in York County.

**Figure 2.**
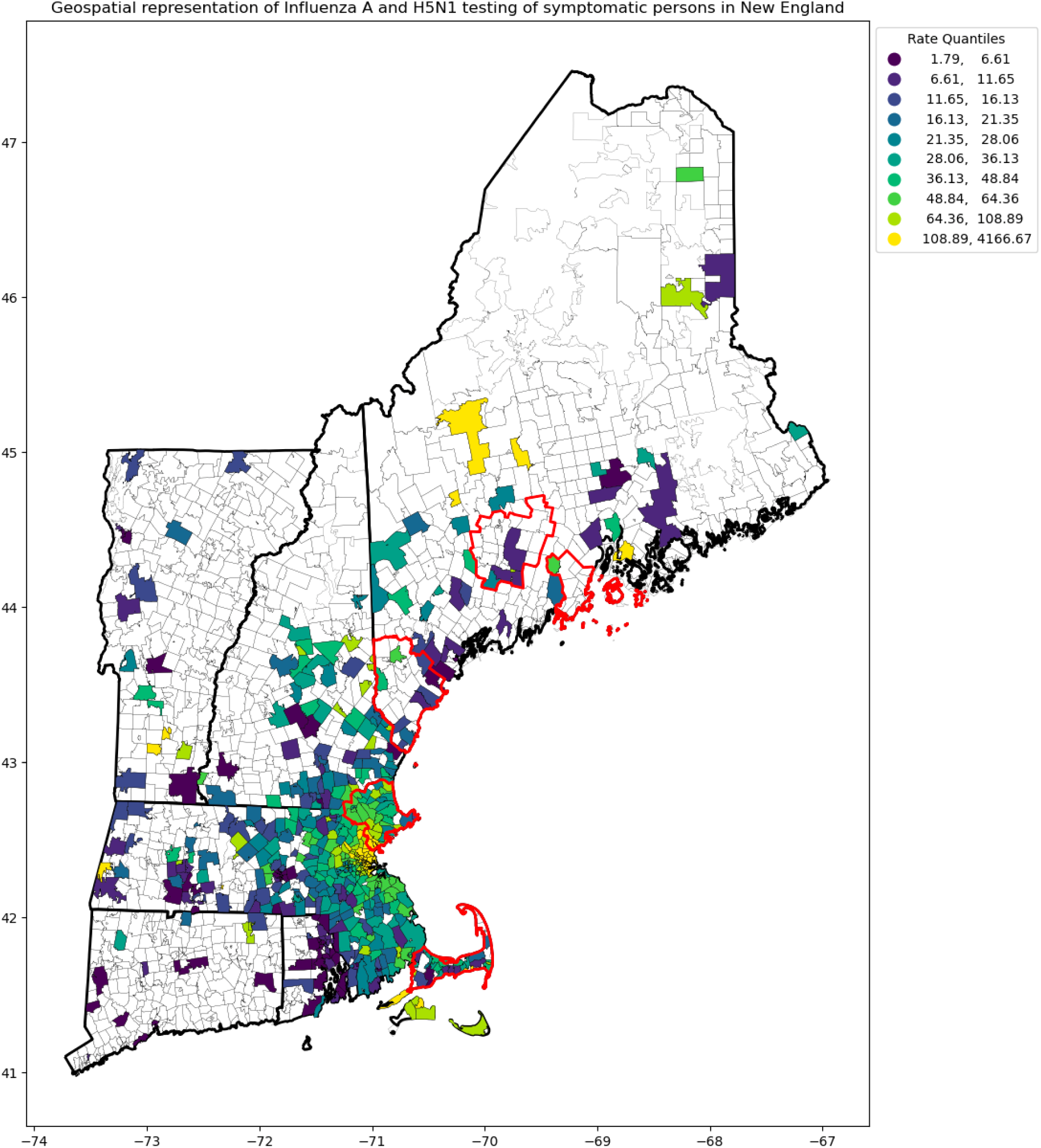
ZIP-level testing rates per 100,000 persons of symptomatic subjects in New England. Data are bucketed into 10 quantiles and color-coded accordingly. State borders are emphasized for visual clarity. Counties in which H5 was confirmed in backyard flocks during the six months preceding the testing window are outlined in red (Barnstable, MA; Essex, MA; York, ME; Kennebec, ME; and Knox, ME). Note that there may be subtle mismatches between the positioning of ZIP and state borders for ZIP codes near state boundaries due to the two different data sources.

### Description of the influenza A-positive cohort

Demographics and clinical features of the 35 influenza A-positive individuals are shown in Figure 3. The median age at the time of specimen collection was 41 years (interquartile range [IQR]: 51 years), including both children and older adults (10/35 [29%] ≤ 15 years old and 7/35 [20%] individuals ≥ 70 years old; Figure 3). All individuals were symptomatic at the time of testing, predominantly with influenza-like illness (ILI) symptoms (31/35 [89%] with cough and 26/35 [74%] with fever). Two individuals (6%) presented with conjunctivitis; conjunctival specimens were not available for H5 testing in these individuals as no H5 risk factors were identified to warrant collection.

**Figure 3.**
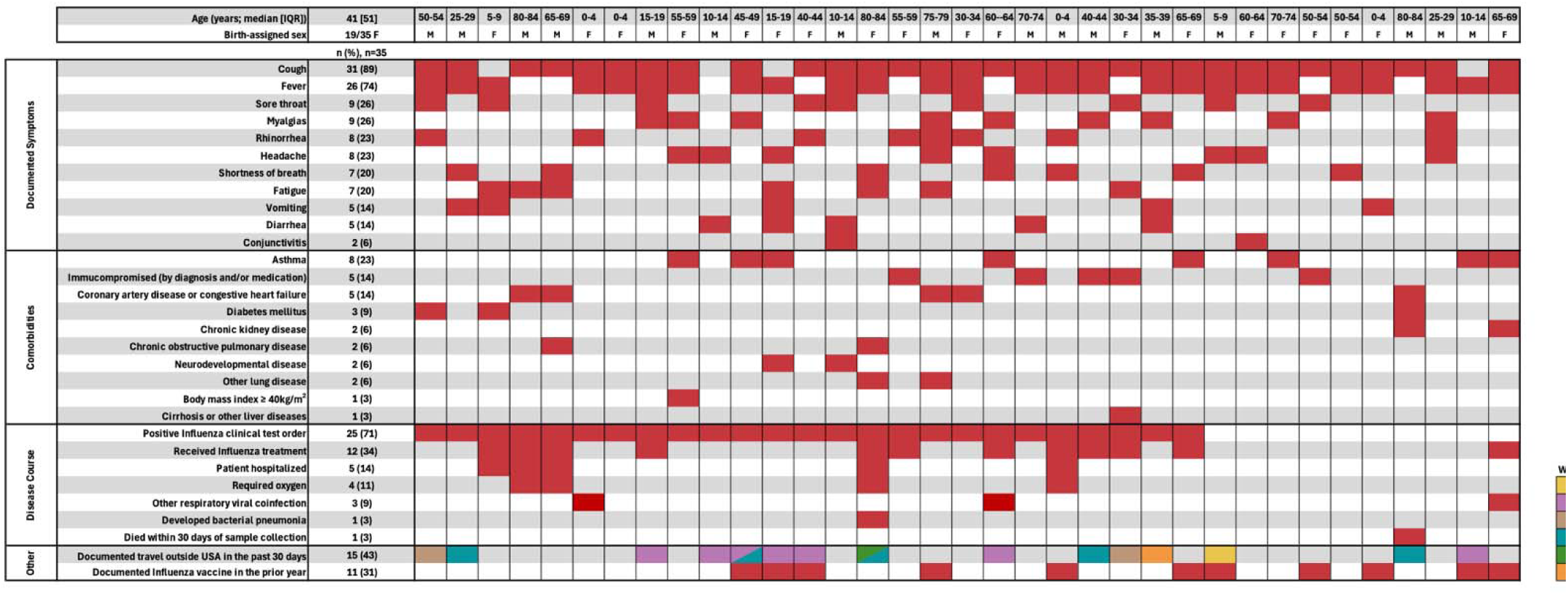
Clinical features, demographics, contextual health information, and other relevant features of Influenza A-positive subjects during the collection period (n=35). All variables were obtained through systematic review of electronic medical records. Documented recent travel history is colored by World Health Organization World Region classification code: African Region (AFR); Region of the Americas (AMR); Eastern Mediterranean Region (EMR); European Region (EUR); South-East Asian Region (SEAR); and Western Pacific Region (WPR; (42). Multiple colors indicate travel to multiple regions within the specified time frame.

Most individuals (22/35 [63%]; Figure 3) had at least one risk factor for severe influenza infection. The most common comorbidities were asthma (8/35, [23%]), immunocompromising conditions due to primary diagnoses or medications (5/35, [14%]), and coronary artery disease or congestive heart failure (5/35, [14%]; see Supplemental Methods). A total of 12/25 (48%) clinically diagnosed individuals received oseltamivir, and 5/35 (14%) subjects required hospitalization. Of the hospitalized subjects, 4/5 (80%) required supplemental oxygen and 2/5 (40%) were admitted to an intensive care unit. Only 3/35 (9%) individuals were found to have viral coinfections (SARS-CoV-2, human rhinovirus/enterovirus, and parainfluenza virus 1). One individual developed bacterial pneumonia, and one died within 30 days of initial influenza A– positive specimen collection, which was not attributed to influenza infection.

Travel outside of the U.S. within 30 days prior to specimen collection was documented for 15/35 (43%) individuals. Most individuals traveled in the Northern Hemisphere; one had returned from equatorial Africa. No individuals had recorded interaction with livestock.

## Discussion

With the emergence of the multistate H5 outbreak in poultry and cattle across the U.S., enhanced surveillance to assess for both zoonotic and human-to-human H5 transmission has been bolstered throughout the country. CDC and some state agencies have recommended testing all individuals with respiratory symptoms for influenza, even in times of low influenza prevalence, to identify obscure H5 cases in the clinical setting (25). Through a research and clinical partnership, a custom-designed algorithm that employed both clinical laboratory and research-developed NAATs was developed that leveraged specimen pooling to screen nearly 7,000 respiratory specimens from close to 6,000 individuals for influenza A and H5 over a two-month period in the summer. Only 35 unique influenza A infections were detected, none of which were identified as H5. These findings align with national surveillance data which have found only three cases of H5 influenza despite testing more than 77,000 specimens (<0.1% detection rate) and provides additional assurance that the risk of zoonotic and human-to-human transmission remains low (26). However, the recent human death from H5, along with widespread disease activity among avian and bovine hosts, underscores the importance of ongoing surveillance (8).

To enhance H5 screening efforts while preserving clinical laboratory resources and ensuring appropriate clinical testing of subjects during a period of low influenza prevalence, FDA guidelines for SARS-CoV-2 NAAT specimen pooling were adapted in the research setting to test for H5 (20,21). This approach was both efficient and effective. Specimen pooling was instrumental for high volume testing while conserving costly clinical laboratory resources, especially when pathogen prevalence was presumed to be low. By testing an additional 2,790 respiratory specimens through research developed protocols, an additional 10 influenza A cases (28.6% of influenza A infections) were detected and ruled out for H5 infection. Elements of this approach—such as leveraging and adapting established clinical workflows through partnership with researchers—may serve as a model for other institutions or regions that wish to conduct surveillance while maintaining guideline-driven clinical testing and preserving clinical laboratory resources.

The testing of nearly 7,000 respiratory specimens during summer of 2024 using a sensitive H5 assay and yielding no cases supports that H5 was unlikely circulating in New England during the study period. Although limited by the single-site nature of our study, previous analyses of respiratory virus activity from a single, major tertiary care center have been generalizable during prior outbreaks, including SARS-CoV-2 and RSV (27–29). Geospatial analysis of the tested individuals, which revealed that 97% and 90% reported residences in New England and Massachusetts, indicated that the catchment area was representative of eastern Massachusetts and, to some extent, across New England, including regions where H5 activity has been observed among avian hosts. However, within the six months prior to initiating H5 testing, H5 was detected in only three backyard flocks in Massachusetts and Maine respectively. No herd or flock outbreaks were reported in the specimen catchment area during the collection period. Without known active viral animal reservoirs in the area during the collection period, the presumed H5 acquisition risk was seemingly low, limiting the utility of performing routine clinical influenza testing in symptomatic individuals as a surrogate for H5 detection during periods of low seasonal prevalence. Future human H5 surveillance efforts may benefit from incorporating active animal or wastewater surveillance data, when available, to guide H5 testing strategies. CDC’s national wastewater surveillance system, which now includes H5 detection from numerous counties across the U.S., can provide an early warning signal for infection at the local level, allowing for rapid implementation of public health measures to prevent spread (30). Data from wastewater surveillance have been used for such purposes in the past, including deployment of poliovirus vaccination campaigns in Israel and India in response to poliovirus detection in sewage (31). The national milk testing strategy that was deployed by USDA in December of 2024 to identify H5-infected herds across the country, is another such strategy that could be used in a similar fashion (32). By maximizing testing from areas where zoonotic transmission is more likely, human cases will likely be identified faster, which would improve implementation of control measures to prevent additional spread and could then be used to direct expanded clinical testing in a data-driven approach.

Given the low incidence of human H5 infection and established risk factors associated with acquisition (exposure to infected animals or contact with contaminated environments), targeted H5 testing based on epidemiology and clinical presentation is an alternative option to large scale surveillance testing when deploying clinical testing resources. With the widespread use of electronic medical records, clinical decision support systems could be implemented to identify potential H5 cases in a systematic manner while preserving laboratory stewardship efforts when overall influenza prevalence is low. Similar tools have been successfully implemented with other emerging infectious diseases, including, Mpox and COVID-19, providing a framework for their development with the H5 outbreak (33,34).

In addition to finding no H5 influenza within our cohort, overall influenza A prevalence was low at only 0.7%, closely aligning with national trends during the same time-period as reported by the CDC National Respiratory and Enteric Virus Surveillance System (NREVSS; 0.7% (35,36)). Most influenza-positive individuals had typical ILI symptoms, including cough (89%) and fever (74%). Hospitalization rates were relatively low (14%) and there was an equal distribution of H3 and H1-2009 subtypes in the strains that underwent partial subtyping (n=26). These data contrast with 2022-2023 national influenza surveillance data in which nearly 65% of influenza A-positive subjects required hospitalization, with H3N2 as the predominant subtype (37). Due to convenience sampling and our small sample size, however, formal conclusions regarding illness severity and strain predominance over the summer months are limited.

Recent international travel was a risk factor for influenza A infection with 15/35 positive individuals (43%) reporting travel within 30 days of test result. This association between travel and influenza infection is consistent with epidemiological data that suggest exposure to new and diverse environments increases the likelihood of contracting respiratory viruses (38,39). Travel history including, locations traveled, itinerary, illness onset in relation to travel, and medications or vaccines received, are key components to a medical evaluation as they help inform the differential diagnosis and determine diagnostic and treatment approaches (40). Our findings support obtaining a travel history as part of routine clinical evaluations with consideration of influenza testing in subjects with respiratory symptoms and recent international travel.

Our study has limitations. First, only convenience respiratory tract samples from symptomatic individuals who sought care within the MGH Clinical Microbiology Laboratory catchment area were included; no non-respiratory samples were collected. Therefore, asymptomatic individuals, those who chose not to be tested, tested at home, or received care elsewhere were not represented. Second, individuals with comorbidities may also be more likely to seek testing, further biasing the specimen collection. Third, although our studies revealed that our pooled strategy did not significantly affect test sensitivity, some sensitivity is inevitably lost with this method, limiting our ability to detect low viral load cases. Finally, regions of New England where livestock are more common and therefore with a higher potential for zoonotic transmission (such as Western Massachusetts) were underrepresented in our cohort (Figure 3). Future surveillance efforts should seek to target these areas specifically as described above, whether through expanded collaborations, outreach programs, or leveraging public health resources and networks.

Additionally, while conjunctivitis is recognized as a defining manifestation of human H5 Influenza infection, no ocular samples were collected by the clinical care teams to assess for influenza RNA presence. Although H5 NAAT from ocular samples in subjects with conjunctivitis has high sensitivity, molecular testing is seldom utilized for diagnosis or treatment of conjunctivitis, and ocular sampling is a nonstandard clinical procedure (41). Nonetheless, obtaining ocular samples from individuals presenting with ILI and ocular symptoms could be a useful tool to increase diagnostic yield when H5 is a clinical concern.

## Conclusion

Implementing a comprehensive summer influenza testing strategy in both clinical and research settings allowed us to identify low-prevalence influenza A cases and confirm the absence of H5 in our catchment area during the study period. Overall, only a small fraction of specimens tested were influenza A-positive, with more than two-thirds of cases identified through routine clinical testing protocols. These findings highlight the need for targeted clinical testing during periods of low influenza prevalence based on epidemiologic risk factors and clinical presentation with enhanced surveillance efforts deployed based on environmental surveillance. Strengthening partnerships between clinical laboratories, public health authorities, infection control, and research teams to help bolster this surveillance will help ensure that any potential H5 circulation is promptly detected while upholding guideline-driven clinical testing and conserving limited clinical resources. With ongoing H5 activity present in animals and humans across the U.S., ongoing H5 surveillance is warranted; through collaboration with the healthcare sector in research capacities, early detection can be bolstered.

## Data Availability

All data produced in the present study are available upon reasonable request to the authors.

## Acknowledgements

The authors thank Paul Biddinger, Jacob Lazarus, and Eileen Searle for their input on this project.

