## Supplemental Material for "Combing the haystacks: The search for highly pathogenic avian influenza virus using a combined clinical and research-developed testing strategy"

**Table of Contents: Supplemental Methods**

**List of Tables and Figures**:

*Pan-influenza A and H5-specific nucleic acid amplification test (NAAT) validation*

We evaluated the overall accuracy of the pan-influenza A and H5-specific NAATs by determining their specificity and sensitivity through both *in silico* and *in vitro* analyses.

For specificity, we performed *in silico* analysis on H5N1 genomes published from the first human cases available as of July 2024 (NCBI BioProject PRJNA1095491; Supplementary Table 6). Multiple sequence alignment was performed using MUSCLE in Geneious Prime (Dotmatics, Boston, MA) with the default parameters. We aligned both primer and probe sets against each published genome, demonstrating 100% nucleotide identity in each primer-probe set (Supplementary Table 6)(1).

To evaluate sensitivity, we performed a modified limit of detection (LOD) analysis by testing serial dilutions of synthetic DNA plasmids designed to include the target regions of the H5 and pan-influenza A assays (sourced from Twist Bioscience, San Francisco, CA; Supplementary Figure 1). The synthetic plasmid for the H5-Specific NAAT had a 301 base pair insert sequence from segment 4 of the *A/goose/Guangdong/1/1996* strain (NCBI RefSeq assembly GCF_000864105.1), nucleotide positions 1,021-1,321. The synthetic plasmid for pan-influenza A NAAT had a 300 base-pair insert that was the consensus sequence of a nucleotide alignment of each segment 7 from the *A/goose/Guangdong/1/1996*, *A/New York/392/2004*, and *A/California/07/2009* strains (NCBI RefSeq assemblies GCF_000864105.1, GCF_000865085.1 and GCF_001343785.1 respectively) spanning nucleotide positions 71-370.

The pan-influenza A NAAT demonstrated an observed sensitivity of at least 1 copy/µL (3/3 replicates positive; Supplementary Table 2, Supplementary Figure 1), while the H5 NAAT showed an observed sensitivity of at least 3.33 copies/µL (3/3 replicates positive; Supplementary Table 2, Supplementary Figure 1). These results are comparable with the reported LOD of other H5 screening NAATs which have reported LODs from <0.5 to 2.5 copies/µL (2).

*In vitro* assay specificity was established using nucleic acid extracted from archived frozen clinical respiratory specimens and H5 RNA derived from milk. The respiratory specimens, which had been tested clinically in the MGH Clinical Microbiology Laboratory, included five H3 influenza A, five H1-2009 influenza A, six influenza B, and 65 pan-negative specimens. These specimens were originally characterized using the BioFire® Respiratory 2.1 (RP2.1) panel (bioMérieux, Salt Lake City, UT)(3). Nucleic acid from H5-positive milk samples was received as extracted eluate from the Sabeti Lab in the Infectious Disease and Microbiome Program at the Broad Institute of MIT and Harvard(4). For clinical specimens, nucleic acid was extracted from 200µL of transport medium from primary samples using the MagMAX Prime Viral/Pathogen Nucleic Acid Isolation kit on the KingFisher Flex platform (ThermoFisher Scientific, Waltham, MA) following the manufacturer's instructions.

Extracted nucleic acid from both clinical specimens and milk underwent quantitative polymerase chain reaction (qPCR) using the pan-influenza A and H5-specific primer/probe sets. Reactions were run in triplicate in a final volume of 20µL per replicate on the QuantStudio™ 6 Pro (ThermoFisher Scientific, Waltham, MA). Each reaction contained 5µL of 4X TaqPath™ 1-Step RT-qPCR Master Mix,  CG (ThermoFisher Scientific, Waltham, MA) 5µL of template nucleic acid, and variable volumes of primers, probes, and nuclease-free water. Final concentrations of primers and probes for each assay are outlined in Supplementary Table 1. Cycling conditions for the reaction were based on recommendations in the Thermofisher TaqPath™ 1-Step RT-qPCR Master Mix, CG User Guide (Supplementary Table 5)(5).

The results of the pan-influenza A and H5-specific NAAT testing are shown in Supplementary Table 3. The pan-influenza A assay correctly detected all influenza A specimens (n=11), including the specimen containing H5 RNA, with no false positive results among influenza-B or pan-negative specimens (n=71). Similarly, the H5-specific assay accurately detected the H5 RNA sample, with no false positive detections in specimens positive for influenza-A H3, H1-2009, influenza B, or pan-negative specimens (n=81). Overall, both assays achieved 100% sensitivity and specificity.

*Modified Pooled Screening Protocol and Validation*

Our pooled screening approach was based primarily on guidance published by the FDA as an emergency use authorization for SARS-CoV-2 screening early in the SARS-CoV-2 pandemic(6). We formed primary biospecimen pools by combining 40 µL of viral transport medium from five randomly selected SARS-CoV-2 negative nasopharyngeal or anterior nasal swab specimens. Each pool was then extracted using the MagMAX Prime Viral/Pathogen Nucleic Acid Isolation kit on the KingFisher Flex platform (ThermoFisher Scientific, Waltham, MA) following the manufacturer's recommendations, and total nucleic acid was eluted in 60µL of provided elution buffer. Each five-specimen pool was tested in triplicate 20µL qPCR reactions as described previously.

To ensure there was no significant loss in sensitivity resulting from the specimen pooling strategy, we compared the average cycle-threshold (CT) change of a positive specimen tested in individual vs. pooled conditions using ten influenza A-positive clinical specimens and 40 influenza A-negative clinical specimens. All pools had one known positive and four known negative specimens. The average CT difference between pooled and individual specimens was 2.3 with a standard deviation of 0.47 (Supplementary Table 4), reflecting an approximate 5-fold difference in RNA levels between pooled and individual specimens. This result is within the 95% confidence interval for the no more than 1.7 CT shift recommended by the FDA EUA pooling guidance(6).

*Clinical and Demographic Data Collection and Analysis*

We gathered clinical and demographic data for individuals who tested positive for influenza through systematic review of electronic medical records by at least two physicians. Information was collected from both structured fields and provider notes using REDCap electronic data capture tools hosted by Mass General Brigham (MGB) Research Computing, Enterprise Research Infrastructure & Services (ERIS) group. REDCap (Research Electronic Data Capture) is a secure, web-based application designed to support data capture for research studies(7).

We defined immunocompromised individuals as having a diagnosis or receiving therapy that is immunosuppressive based on internal MGB guidelines(8). Diagnoses considered included active lymphoma or leukemia (including indolent chronic lymphocytic leukemia), metastatic cancer (unless the disease had been stable for at least one year), aplastic anemia, congenital immunodeficiency, solid organ transplant (with immunosuppressive therapy), hematopoietic stem cell transplant (unless more than two years post-transplant), or having undergone cytotoxic chemotherapy within the preceding three months. Immunosuppressive therapies considered included glucocorticoid therapy equivalent to ≥20 mg/day of prednisone for ≥2 weeks (or discontinued within the past month), alkylating agents or antimetabolites (methotrexate >0.4 mg/kg/week, azathioprine >3 mg/kg/day, 6-MP >1.5 mg/kg/day) within the past three months, calcineurin inhibitors (e.g., cyclosporine, tacrolimus), mTOR inhibitors (e.g., sirolimus, everolimus), mycophenolate mofetil, or biologic immunosuppressants/immunomodulators (excluding immune checkpoint inhibitors) within the past three months (six months for lymphocyte-depleting agents). Immunocompromised individuals in this cohort had diagnoses of either metastatic cancer or active lymphoma/leukemia or were receiving biologic immunosuppressants or alkylating agents within the last three months.

We documented travel outside of the United States for 15 of the 35 influenza-positive individuals. Specific countries visited included Bermuda, Canada, the Dominican Republic, El Salvador, Iceland, India, Pakistan, Portugal, Uganda, the United Arab Emirates, and Vietnam

Figures and tables:

| Target Name | Sequence Description | Oligonucleotide Sequence (5' to 3') | Final Concentrations ($\mu$L) |
|---|---|---|---|
| Pan-Influenza A NAAT | Forward 1 | CAA GAC CAA TCY TGT CAC CTC TGA C | 0.4 |
| | Forward 2 | CAA GAC CAA TYC TGT CAC CTY TGA C | 0.4 |
| | Reverse 1 | GCA TTY TGG ACA AAV CGT CTA CG | 0.6 |
| | Reverse 2 (MGH) | GCA **Y**TT TGG A**Y**A AAG CGT CTA CG | 0.8 |
| | Probe | FAM/TGC AGT CCT/ZEN/CGC TCA CTG GGC ACG/3IABkFQ | 0.2 |
| H5-Specific NAAT | Forward | TAT AGA RGG AGG ATG GCA GG | 0.8 |
| | Reverse | ACD GCC TCA AAY TGA GTG TT | 0.8 |
| | Probe | FAM/AGG GGA GTG/ZEN/GKT ACG CTG CRG AC/3IABkFQ | 0.2 |
| Rnase P | Forward | AGA TTT GGA CCT GCG AGC G | 0.8 |
| | Reverse | GAG CGG CTG TCT CCA CAA GT | 0.8 |
| | Probe | Cy5/TTC TGA CCT/TAO/GAA GGC TCT GCG CG/3IAbRQSp | 0.2 |

*Supplementary Table 1*. Primer and probe sequences used for the Pan-influenza A NAAT, H5-Specific NAAT and RNase P internal positive control. The Pan-Influenza A reverse primer 2 was edited to be the reverse 2 (MGH) primer. Areas where the primer sequence was edited are bolded and underlined, and both changes were from a T base to a Y degenerate base. Final concentrations used for RT-qPCR are from the CDC with the exception of the Reverse 2 (MGH) primer(9). The Pan-Influenza A Forward 1, Forward 2, Reverse 1, probe and the entire RNase P primer-probe set are from the CDC Flu Sars-Cov-2 Multiplex assay. The H5-Specific NAAT primer and probe set were also published previously(10).

NAAT: nucleic acid amplification test; MGH: Massachusetts General Hospital

| **Pan-Influenza A NAAT** | | | | | | |
|---|---|---|---|---|---|---|
| copies/μL | 1 | 3.33 | 10 | 100 | 1000 | 10,000 |
| % positive (out of 3) | 100% | 100% | 100% | 100% | 100% | 100% |
| Mean Ct (SD) | 36.456 (1.852) | 34.823 (0.745) | 32.194 (0.481) | 29.145 (0.150) | 25.685 (0.099) | |
| **H5-Specific NAAT** | | | | | | |
| copies/μL | 1 | 3.33 | 10 | 100 | 1000 | 10,000 |
| % positive (out of 3) | - | 100% | 100% | 100% | 100% | 100% |
| Mean Ct (SD) | - | 34.067 (1.487) | 32.715 (0.252) | 29.6 (0.058) | 26.298 (0.009) | |

*Supplementary Table 2*. Modified limit of detection (LOD) analysis of serial dilutions of quantified synthetic Pan-influenza A and H5 DNA plasmids on the Pan-influenza A and H5-specific NAATs. Observed pan-influenza A NAAT LOD was ≤ 1 copy/µL (positive in 3/3 replicates). Observed H5 NAAT LOD was ≤ 3.33 copies/µL (positive in 3/3 replicates).

Rxn: reaction; Ct: cycle-threshold; SD: standard deviation.

| Clinical Specimens (n=81) | INFA (%) | H5 (%) |
|---|---|---|
| H5 (1)[1] | 1/1 (100) | 1/1 (100) |
| H3 (5) | 5/5 (100) | 0/5 (0) |
| H1-2009 (5) | 5/5 (100) | 0/5 (0) |
| Influenza B (6) | 0/6 (0) | 0/6 (0) |
| pan-negative (65) | 0/65 (0) | 0/65 (0) |

*Supplementary Table 3*. *In vitro* accuracy testing of the Pan-influenza A (INFA) and H5-Specific (H5) NAATs using clinical specimens and H5 RNA derived from milk. Clinical specimens were tested using the BioFire Respiratory 2.1 Panel which includes the following targets: SARS-CoV-2, non-SARS-CoV-2 coronaviruses, human rhinovirus/enterovirus (combined target), influenza A including H1, H3, and H1-2009 subtypes), influenza B, respiratory syncytial virus, parainfluenza viruses 1-4, adenovirus, human metapneumovirus, *Bordetella pertussis* and *parapertussis, Mycoplasma pneumoniae,* and *Chlamydia pneumoniae.*

[1]Represents H5 RNA derived from milk from H5 positive cattle.

NAAT: nucleic acid amplification test; RNA: ribonucleic acid.

| Specimen | CT change ± SD |
|---|---|
| Sample 1 H1-2009 | 1.9 ± 0.12 |
| Sample 2 H1-2009 | 2.6 ± 0.1 |
| Sample 3 H1-2009 | 2.9 ± 0.26 |
| Sample 4 H1-2009 | 2.3 ± 0.12 |
| Sample 5 H1-2009 | 1.8 ± 0.3 |
| Sample 1 H3 | 2.4 ± 0.08 |
| Sample 2 H3 | 1.8 ± 1.31 |
| Sample 3 H3 | 2.8 ± 0.41 |
| Sample 4 H3 | 2 ± 0.34 |
| Sample 5 H3 | 2.8 ± 0.19 |
| average CT change ± SD | 2.3 ± 0.47 |
| CT 95% CI ± SD | 2.3 ± 0.94 |

*Supplementary Table 4*. Average cycle-threshold (CT) change of influenza A positive specimens tested in individual vs. pooled conditions. Pooled specimens included one influenza A positive specimen combined with four influenza A negative specimens. SD represents standard deviation; CI represents confidence interval and CT represents cycle threshold.

| Step | Temperature | Time | Cycles |
|---|---|---|---|
| UNG incubation | 25°C | 2 minutes | 1 |
| Reverse transcription | 55°C | 15 minutes | 1 |
| Polymerase activation | 95°C | 2 minutes | 1 |
| Amplification | 95°C | 3 Seconds | 40 |
| | 60°C | 30 Seconds | |

*Supplementary Table 5:* The cycling conditions used to perform the Pan-influenza A and H5-specific NAAT on the QuantStudio™ 6 Pro (ThermoFisher Scientific, Waltham, MA).

PCR: polymerase chain reaction.

| Genome ID | State of Origin | Release date | H5-Specific NAAT % alignment | Pan-Influenza NAAT % alignment |
|---|---|---|---|---|
| GCA_039465435.1 | Texas | April, 2024 | 100% | 100% |
| GCA_039991525.1 | Michigan | May, 2024 | 100% | 100% |
| GCA_040996195.1 | Colorado | Juy, 2024 | 100% | 100% |
| GCA_040780915.1 | Colorado | Juy, 2024 | 100% | 100% |
| GCA_040995305.1 | Colorado | Juy, 2024 | 100% | 100% |
| GCA_040995325.1 | Colorado | Juy, 2024 | 100% | 100% |

*Supplementary Table 6: In silico* analysis in Geneious prime of the pan-influenza A NAAT and the H5-Specific NAAT against the six H5N1 human genomes as of July 2024. Alignments allowed 0 mismatches; sequences are from NCBI BioProject PRJNA1095491.

| **ZIP** | **City** | **County** | **State** |
|---|---|---|---|
| 01199 | Springfield | Hampden | Massachusetts |
| 01831 | Haverhill | Essex | Massachusetts |
| 01842 | Lawrence | Essex | Massachusetts |
| 01885 | West Boxford | Essex | Massachusetts |
| 01888 | Woburn | Middlesex | Massachusetts |
| 02051 | Marshfield | Plymouth | Massachusetts |
| 02059 | Marshfield | Plymouth | Massachusetts |
| 02153 | Medford | Middlesex | Massachusetts |
| 02196 | Boston | Suffolk | Massachusetts |
| 02205 | Boston | Suffolk | Massachusetts |
| 02238 | Cambridge | Middlesex | Massachusetts |
| 02303 | Brockton | Plymouth | Massachusetts |
| 02327 | Pembroke | Plymouth | Massachusetts |
| 02334 | Easton | Bristol | Massachusetts |
| 02345 | Manomet | Plymouth | Massachusetts |
| 02361 | Plymouth Center | Plymouth | Massachusetts |
| 03821 | Dover | Strafford | New Hampshire |
| 10163 | New York | New York | New York |
| 10179 | New York | New York | New York |
| 32969 | Vero Beach | Indian Riber | Florida |
| 34611 | Spring Hill | Hernando | Florida |

*Supplementary Table 7*. Manually populated ZIP Code information. Data are obtained from various sources including: UnitedStatesZipCodes, MapQuest, and ZipDataMaps(11–13).

| Confirmation Date | State | County Name | Production | Birds Affected |
|---|---|---|---|---|
| 18th March 2024 | Maine | Knox | WOAH Non-Poultry | 40 |
| 7th March 2024 | Massachusetts | Essex | WOAH Poultry | 70 |
| 6th February 2024 | Massachusetts | Essex | WOAH Non-Poultry | 20 |
| 25th January 2024 | Maine | Kennebec | WOAH Non-Poultry | 40 |
| 16th January 2024 | Massachusetts | Barnstable | WOAH Non-Poultry | 150 |
| 10th January 2024 | Maine | York | WOAH Non-Poultry | 60 |

*Supplementary Table 8*. Flock outbreaks in New England during the 6-month period before the testing window (June 23rd – August 28th 2024), according to the Animal and Plant Health Inspection Service of the U.S. Department of Agriculture(14).

| State | Count | Latest H5 Flock Outbreak | Flock Outbreaks During Testing Window |
|---|---|---|---|
| Florida | 44 | 14th January 2025 | 4 (7/19, 7/24, 8/5, 8/20) |
| New York | 32 | 26th February 2024 | - |
| California | 15 | 3rd January 2025 | - |
| New Jersey | 10 | 15th September 2023 | - |
| Texas | 7 | 9th January 2025 | - |
| Maryland | 7 | 14th January 2025 | - |
| North Carolina | 6 | 7th January 2025 | - |

*Supplementary Table 9.* Testing counts and flock outbreaks in Non-New England states in which more than 5 persons were tested during the testing window (June 23rd – August 28th 2024)(14).

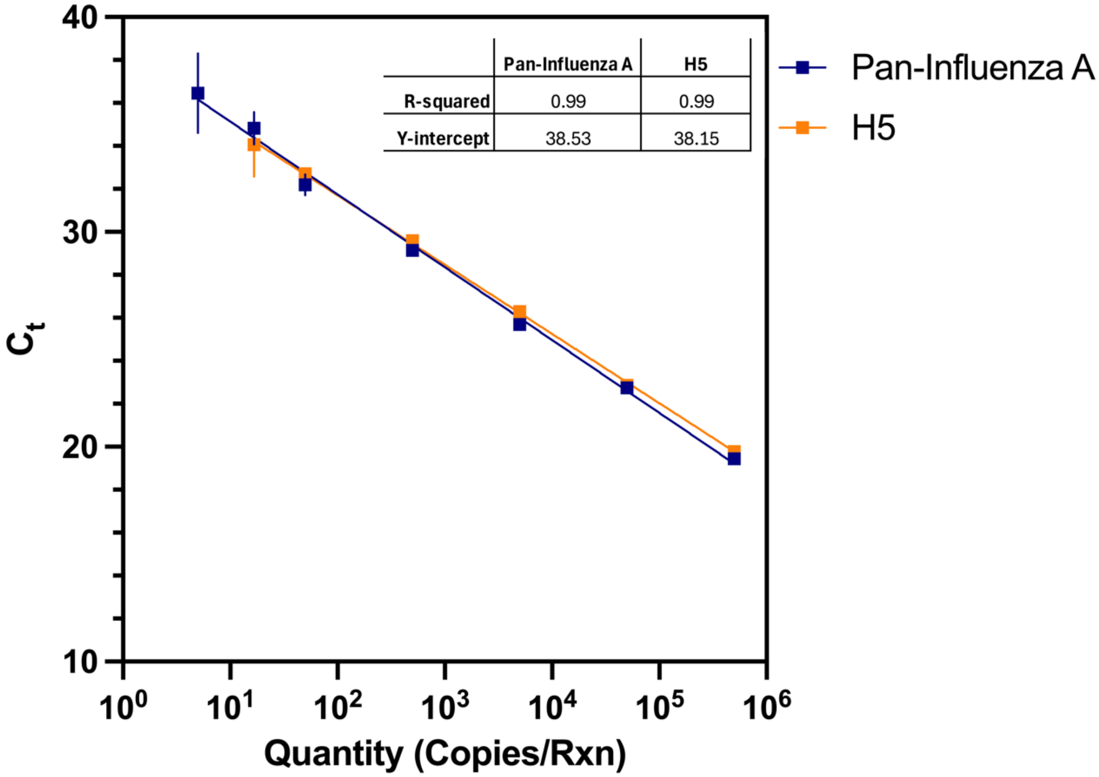

| | Pan-Influenza A | H5 |
|---|---|---|
| R-squared | 0.99 | 0.99 |
| Y-intercept | 38.53 | 38.15 |

*Supplementary Figure 1*. Standard curves generated by performing serial dilutions of quantified synthetic pan-influenza A and H5-specific DNA plasmids on the Pan-Influenza A and H5-specific (H5) NAAT. All reactions were run in triplicate. For the pan-influenza NAAT, observed limit of detection (LOD) was at least 5 copies/reaction; for the H5 NAAT, LOD was at least 16.65 copies/reaction. Error bars represent one standard deviation from the mean.

NAAT: nucleic acid amplification test; CT: cycle-threshold.
